# Childhood Maltreatment Causally Associated with Osteoarthritis: A Mendelian Randomization Study

**DOI:** 10.1101/2025.11.25.25340987

**Authors:** Zekai Yu, Jianyu Zhu

**Author notes:** **Corresponding authors**: Jianyu Zhu.

## Abstract

**Background:** To explore the causal relationship between childhood maltreatment (CM) and osteoarthritis (OA) and identify mediating factors such as major depressive disorder (MDD), body mass index (BMI), alcohol intake frequency, educational attainment, and smoking initiation.

**Methods:** Univariable and multivariable Mendelian randomization (MR) analyses were conducted to assess the causal associations and the mediating effects of the specified risk factors.

**Results:** The analyses provided robust evidence supporting a causal association between CM and an increased risk of OA (OR=1.682, CI=1.414-2.001). Sensitivity analyses confirmed the consistency of these associations, with no significant outliers detected. Mediation analysis showed that BMI accounted for 13.46% of the CM-OA association (beta indirect association=0.070, CI=0.030-0.089), MDD for 8.27% (beta indirect association=0.043, CI=0.033-0.140), smoking initiation for 11.73% (beta indirect association=0.061, CI=0.017-0.063), alcohol intake frequency for 30.00% (beta indirect association=0.156, CI=0.016-0.062), and educational attainment for 26.23% (beta indirect association=0.132, CI=0.013-0.054).

**Conclusions:** Our findings suggest a genetically supported causal relationship between CM and OA, with BMI, MDD, alcohol intake frequency, educational attainment, and smoking initiation serving as significant mediators.

## Introduction

Childhood maltreatment (CM) is a complex global health crisis affecting up to 36% of the population^(1)^, significantly impacting individual and societal well-being. It is estimated that one in five women and one in thirteen men have experienced sexual abuse^(2)^, while nearly a quarter of adults have endured physical abuse during their childhood^(2)^. In most cases, family members, including parents, are responsible for up to 80% of these incidents^(3, 4)^.

CM could be intergenerational transmitted due to the interplay of social factors and genetics^(5)^. Genetic studies show that CM has a heritability of 6-62%^(6)^. Additionally, a study in 2020 found that single nucleotide polymorphisms (SNPs) contribute to 6% of this heritability^(7)^. These findings underscore the importance of considering genetic influences when examining the long-term health impacts of CM.

Osteoarthritis (OA) is a prevalent chronic musculoskeletal disease, that mainly affects the knees and hips. OA is characterized by cartilage degradation, subchondral bone remodeling, and synovitis^(8, 9)^. OA affects over 500 million people globally ^(10)^. The prevalence of OA is still increasing due to the population aging and obesity^(11, 12)^. Additionally, OA places a significant burden on public healthcare systems, often necessitating joint replacement surgery in severe cases^(13, 14)^. OA is a complex disease with high heterogeneity caused by the interplay of genetic, metabolic, biomechanical, and environmental influences^(15–18)^. An increasing number of studies have consistently demonstrated a potential relationship between CM and OA^(19–21)^.

The causal relationship between CM and OA, as well as the mediated factors, remain not very clear. Cross-sectional designs were the most common protocol used in previous studies, which could not strictly eliminate confounding factors. CM is closely associated with the risk of increased body mass index (BMI)^(3, 22)^, smoking^(4, 23)^, depression^(24, 25)^, alcohol use^(3, 4, 26)^, and educational attainment^(24, 27)^. These risk factors could also play a role in the development of OA. We hypothesized that suggesting that BMI, smoking, depression, alcohol use, and educational attainment might mediate the relationship between CM and OA. However, self-reported measures for both CM and OA introduce biases due to the subjective nature of these reports, complicating the interpretation of the data^(28)^. Clarifying the causal association between CM and OA is important for effective health interventions and social support.

Ethical and practical issues of conducting randomized trials pose a barrier to exploring the causal relationship between CM and OA. Mendelian randomization (MR) offers a robust alternative to observational studies by utilizing genetic variants as instrumental variables (IVs) to estimate the causal effects of exposures on outcomes, thereby mitigating common confounders^(29)^. In studying the impact of CM on OA, MR allows researchers to sidestep issues such as confounding factors, reverse causation, and measurement errors that often plague conventional observational research. This method leverages the random assortment of genes from parents to offspring during conception, which ensures that the genetic variation associated with CM is independent of the confounding variables that typically affect observational assessments. The validity of MR depends on three key assumptions: (a) IVs must be strongly associated with the exposure^(29)^, (b) IVs should not be linked to any confounders^(29)^, and (c) IVs can only affect the outcome through the exposure, not by any direct or indirect paths^(29)^.

In this study, the primary objective is to utilize data from large-scale genome-wide association studies (GWAS) to evaluate the causal association between CM and OA using MR. The secondary objective is to perform multivariable MR (MVMR) and mediation MR analysis to identify confounders and determine to what extent that BMI, smoking, and alcohol explain the association between CM and OA.

## Materials and Methods

### The source of GWAS summary data

In this study, we utilized summary statistics from comprehensive GWAS datasets exclusively comprising individuals of European ancestry (Table 1). The extensive availability of GWAS data from European cohorts provided a substantial dataset, enhancing the robustness of our analysis. Additionally, focusing exclusively on this homogeneous population minimized the risk of population stratification, which can distort genetic associations. This careful selection ensures the accuracy of our causal inferences by preserving uniform genetic architecture.

**Table 1.** Characteristics of all summary genome-wide association studies conducted in our MR study.

| Role | Phenotype | Accessed ID | Dataset source | Sample size | Cases |
| --- | --- | --- | --- | --- | --- |
| Exposure | Childhood maltreatment<sup>(6)</sup> | - | Apollo | 185,414 | - |
| Outcome<br>(discovery) | Osteoarthritis<sup>(30)</sup> | - | GO | 826,690 | 177,517 |
| Outcome<br>(replication) | Osteoarthritis<sup>(31)</sup> | ukb-b-14486 | MRC-IEU | 462,933 | 38,472 |
| Mediator | Body mass index (BMI)<sup>(31)</sup> | ukb-b-19953 | MRC-IEU | 461,460 | - |
|  | Smoking initiation<sup>(32)</sup> | ieu-b-4877 | MRC-IEU | 607,291 | 311,629 |
|  | Major depressive disorder (MDD)<sup>(33)</sup> | ieu-a-1188 | MRC-IEU | 173,005 | 59,851 |
|  | Alcohol intake frequency<sup>(31)</sup> | ukb-b-5779 | MRC-IEU | 462,346 | - |
|  | Educational attainment<sup>(34)</sup> | ebi-a-GCST90<br>029013 | MRC-IEU | 461,457 | - |

Abbreviations: MR: Mendelian randomization; Apollo: University of Cambridge Repository; GO: Genetics of Osteoarthritis (GO) Consortium; MRC-IEU: MRC Integrative Epidemiology Unit;

#### Exposure

Childhood maltreatment summary statistics were derived from Warrier’s study^(6)^, which incorporated GWAS summary statistics from the Psychiatric Genomics Consortium (PGC; n = 26,290) and individual-level data from the UK Biobank (UKBB; n = 143,473), Avon Longitudinal Study of Parents and Children (ALSPAC; n = 8,346), Adolescent Brain Cognitive Development Study (ABCD; n = 5,400), and Generation R (GenR; n = 1,905). Childhood maltreatment encompassed emotional and physical neglect as well as emotional, physical, and sexual abuse, with prospective reports largely provided by parents or caregivers and retrospective accounts self-reported. The participants’ ages ranged from 10 to 64 years. Association testing across the UKBB, ALSPAC, and ABCD utilized linear mixed-effects models to adjust for relatedness and population stratification, while the UKBB and ABCD also incorporated 20 genetic principal components to further refine controls for stratification. In GenR, linear regression was applied for association analyses, including five principal components as covariates to ensure rigorous adjustment for potential confounders.

#### Outcome

For the discovery phase, the summary datasets related to All Osteoarthritis (All OA) were derived from a GWAS meta-analysis that included 826,690 individuals across nine distinct populations, comprising 177,517 OA patients and 649,173 controls^(30)^. OA cases were sourced from the UK Biobank and Arthritis Research UK Osteoarthritis Genetics (arcOGEN) databases. Controls were obtained from the United Kingdom Household Longitudinal Study (UKHLS), a longitudinal panel survey encompassing 40,000 UK households across England, Scotland, Wales, and Northern Ireland. Osteoarthritis was defined using multiple criteria including self-reported OA, clinical diagnoses, ICD-10 codes, or radiographic evidence, depending on the data available in each cohort. Controls were either OA-free or population-based, with some cohorts including or excluding participants based on ICD codes. All OA encompassed osteoarthritis at any of the investigated joint sites: hip, knee, hand, finger, thumb, and spine.

For the replication phase, osteoarthritis dataset were obtained from a publicly available GWAS dataset (Accessed ID: ukb-b-14486) ^(31)^. This dataset, constructed by the MRC Integrative Epidemiology Unit (MRC-IEU) consortium using the UK Biobank, included 462,933 Europeans, comprising 38,472 cases and 424,461 controls.

#### Mediator

The body mass index (BMI) GWAS data (Accessed ID: ukb-b-19953) included 461,460 Europeans from the UK Biobank. The smoking initiation GWAS data (Accessed ID: ieu-b-4877) included 607,291 Europeans (311,629 cases and 295,662 controls) from the GWAS & Sequencing Consortium of Alcohol and Nicotine use (GSCAN) ^(32)^. The major depressive disorder (MDD) GWAS data (Accessed ID: ieu-a-1188) included 173,005 Europeans (59,851 cases and 113,154 controls) from the Psychiatric Genomics Consortium (PGC) ^(33)^. The Alcohol intake frequency GWAS data (Accessed ID: ukb-b-5779) included 462,346 Europeans from the UK Biobank^(31)^. The Educational attainment GWAS data (Accessed ID: ebi-a-GCST90029013) included 461,457 Europeans from Loh ’s study^(34)^. All datasets were provided by the MRC Integrative Epidemiology Unit (MRC-IEU) consortium^(31)^.

## Statistical analysis

The overview for our MR analysis was illustrated in Figure 1. We conducted all statistical computations through the R computing environment (version 4.3.0). For the purposes of MR and additional sensitivity assessments, we utilized the analytical capacities of the R packages TwoSampleMR (version 0.6.2), MR-PRESSO (version 1.0.0), MRcML (version 0.0.0.9000), and MVMR (version 0.4). We strictly followed the guidelines of STROBE-MR for reporting the outcomes of our MR analysis.

**Figure 1:**
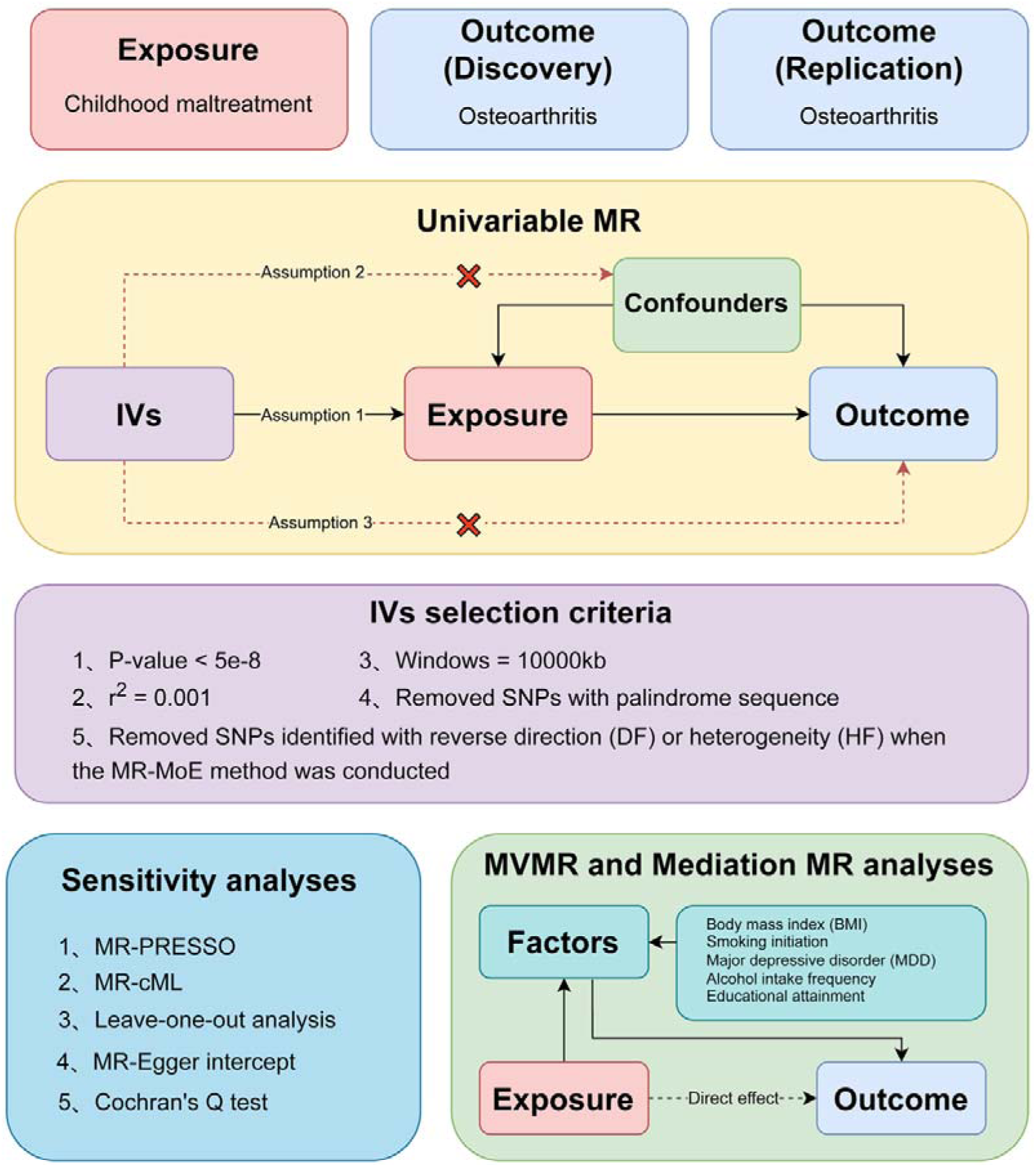
Overview of the study.

### TSMR (Two-Sample Mendelian randomization)

#### IVs selection

First, we implemented a rigorous significance threshold of P-value < 5e-8 for CM to ensure their reliability. Second, to mitigate the impact of linkage disequilibrium, we utilized an LD parameter (r^2^) cutoff of 0.001 and a genetic distance of 10,000 kilobases, referencing the European (EUR) 1000 Genomes Project Phase 3 dataset^(35)^. Third, our selection process required a minimum F-statistic of 10, signifying the robustness of the IVs. Fourth, we excluded IVs directly associated with the outcome to maintain their independence, using a P-value threshold of 5e-8.

#### Discovery MR analysis

The analysis consists of two steps. First, we perform a univariable MR (UVMR) analysis to evaluate the causal association between CM and OA. Second, we conduct complementary sensitivity analysis.

We employed the Mendelian Randomization-Mixture of Experts (MR-MoE) method to evaluate the causal association between CM and OA^(36)^. The MR-MoE method, which integrates traditional Mendelian Randomization with a mixture of expert models, offers several advantages in addressing pleiotropy and heterogeneity in genetic instruments, thereby improving the robustness of causal inference^(36)^.The results for MR-MoE rename the methods to be the estimating method plus the instrument selection method. There are four instrument selection methods: Tophits (no filtering), directional filtering (DF, a non-thresholded adaptation of Steiger filtering), heterogeneity filtering (HF, excluding instruments that significantly impact Cochran’s Q statistic with p < 0.05), and a combined approach (DF + HF) where DF is applied first, followed by HF^(36)^. Furthermore, MR-MoE adds the MOE column, which is a predictor for each method’s performance in terms of high power and low type 1 error, scaled from 0 to 1, where 1 indicates the best performance^(36)^. We selected the method with the highest MOE predictor to determine the most reliable MR estimate.

In addition, we performed three complementary sensitivity analyses to detect and adjust for horizontal pleiotropy and heterogeneity. The MR-PRESSO method was utilized to identify and correct for horizontal pleiotropy by removing significant outliers^(37)^. Specifically, the MR-PRESSO global test was applied to assess the presence of horizontal pleiotropy, with a P-value greater than 0.05 indicating a low likelihood of horizontal pleiotropy. The MR-PRESSO outlier test, which mandates a minimum of 50% genetic variation for validity, operates under the InSIDE (Instrument Strength Independent of Direct Effect) assumption^(37)^. To address pleiotropy that is both correlated and uncorrelated, we applied the constrained cML-MA, which does not depend on the InSIDE assumption^(38)^. This method provides robust control over both related and unrelated pleiotropic effects^(38)^. Lastly, a leave-one-out (LOO) analysis was conducted to evaluate the impact of individual SNPs on the overall results, ensuring robustness and reliability^(39)^

#### Replication MR analysis

To validate the robustness of our discovery phase MR results, we conducted a replication MR analysis. We considered the association between CM and OA to be reliable if the P-value <0.05 obtained from the MR-MoE method.

### MVMR and Mediation MR

To distinguish between mediators and confounders in the causal relationship between childhood maltreatment (CM) and osteoarthritis (OA), we employed multivariable Mendelian randomization (MVMR) and Mediation MR analyses. MVMR enabled us to disentangle the roles of body mass index (BMI), smoking initiation, major depressive disorder (MDD), alcohol intake frequency, and educational attainment, determining whether these factors act as mediators or confounders in the association between CM and OA. By adjusting for these factors, we assessed if CM, as an exposure, remained significantly associated with OA. A continued significant association would suggest that these factors are more likely to be mediators rather than confounders. This analysis also provided the direct causal effect of CM on OA.

In cases where these factors were identified as mediators, they were considered part of the causal pathway from CM to OA. This was further explored through mediation MR analysis, which quantified the mediation effect of CM on OA via these mediators. Specifically, we conducted MR-MoE to examine whether there were causal effects in both the CM to mediator and mediator to OA pathways. The total association (total effect) was derived from the MR analyses mentioned earlier. The mediation effect was calculated by subtracting the direct effect from the total effect, and the proportion mediated was determined by dividing the mediation effect by the total effect. Notably, the mediation analyses were conducted using the FE IVW - DF+HF method from the MR-MoE analyses to obtain robust results.

## Results

### Instrumental variables

We identified 10 independent SNPs associated with CM with a P-value < 5e-8 to be used as instrumental variables. After excluding rs12031035 due to its palindromic nature with intermediate allele frequencies, we utilized a total of 9 SNPs in the MR-MoE analyses for OA. These SNPs exhibited strong instrumental variable characteristics, with a mean F-statistic of 36.505 (median 33.454, range 29.877-55.338), indicating their robustness and suitability for the analysis (Supplementary material Table S1). Additionally, the MR-MoE method detected heterogeneity in rs1859100, rs35560901, and rs77987546, but no SNPs were identified as having a potential for being directional filtered.

### Casual association between CM and OA

We found evidence for an association between CM and OA (Figure 2). In the discovery phase, we initially assessed the association between CM and OA using the IVW no filtering method (IVW - Tophits), revealing a significant relationship (P_FE IVW_=1.21E-08; P_RE IVW_=0.00839). To obtain a more precise estimate, we employed the Fixed Effects Inverse Variance Weighted method excluding SNPs filtered for heterogeneity (FE IVW - HF) since it is the highest scoring moe predictor method. This approach yielded an odds ratio (OR) of 1.682 (95% CI: 1.414-2.001, P = 4.24E-09), indicating a 68% increased odds of OA per unit increase on the CM scale. Out of nine SNPs considered as instrumental variables, six were utilized, while three were excluded due to heterogeneity.

**Figure 2:**
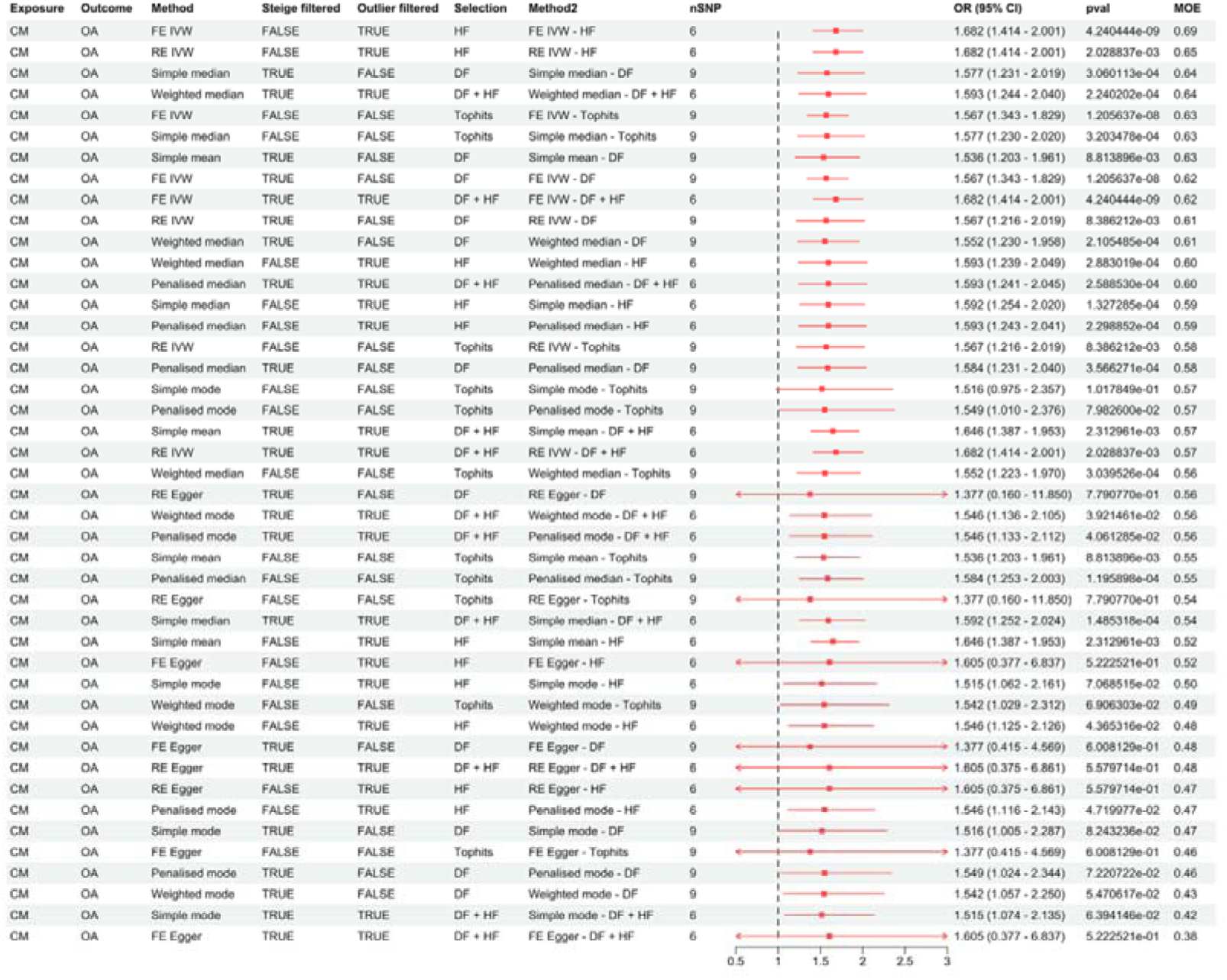
Forest plot depicting the MR-MoE analysis results for CM and OA. Abbreviations: FE IVW: Fix Effects Inverse Variance Weighting; RE IVW: Random Effects Inverse Variance Weighting; Tophits: No Filtering; DF: Directional Filtering; HF: Heterogeneity Filtering; CI: Confidence Interval; OR: Odds Ratio

Using the FE IVW - HF method, Cochran’s Q test confirmed no heterogeneity in the association between CM and OA (Q = 3.96; DF = 5; P = 0.555). The Fixed Effect Egger intercept showed no evidence of unbalanced horizontal pleiotropy (beta = 0.196; SE = 3.06; P = 0.952). Additionally, the cML-MA method did not identify pleiotropic effects, and the association remained significant (beta = 0.529; SE = 0.101; P = 1.447124e-07). The MR-PRESSO global test did not detect any outliers (P_global test_ = 0.571), and the leave-one-out analysis confirmed the absence of outlier SNPs (Figure 3).

**Figure 3:**
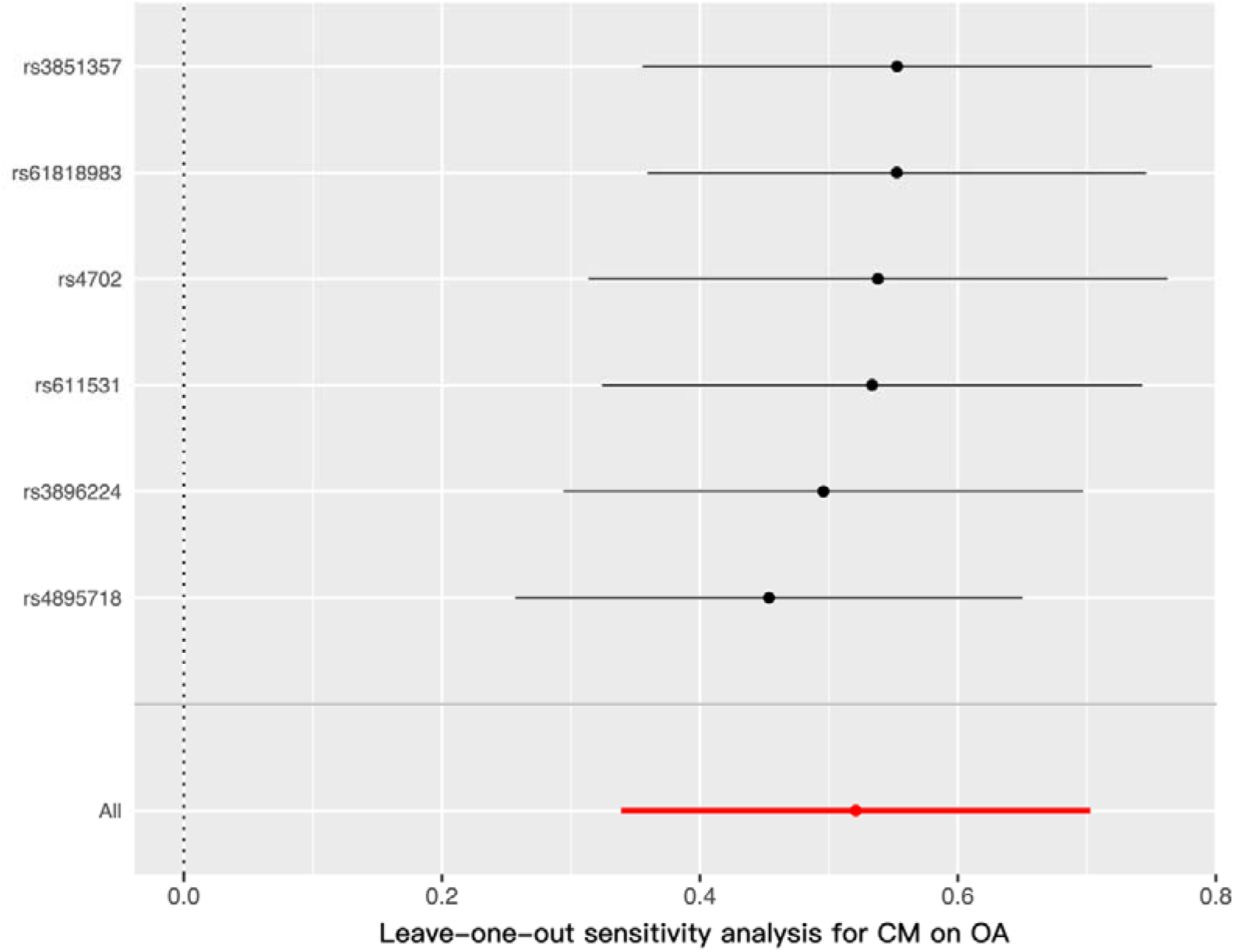
Re-evaluating the association between CM and OA by sequentially excluding individual SNP. Note: The y-axis represents the SNP that was excluded. Excluding any of the SNPs does not render the association non-significant.

During the replication phase, MR results confirmed a causal relationship between CM and OA. The MR-MoE method was used for robust estimation, with detailed results available in (Supplementary material Table S2).

### MVMR and Mediation MR

#### MVMR

MVMR analysis indicate that CM remains significantly associated with OA after adjusting for potential confounders such as BMI, MDD, smoking, alcohol and education. These findings suggest that the relationship between CM and OA is robust and persists even when accounting for these factors (Figure 4).

**Figure 4:**
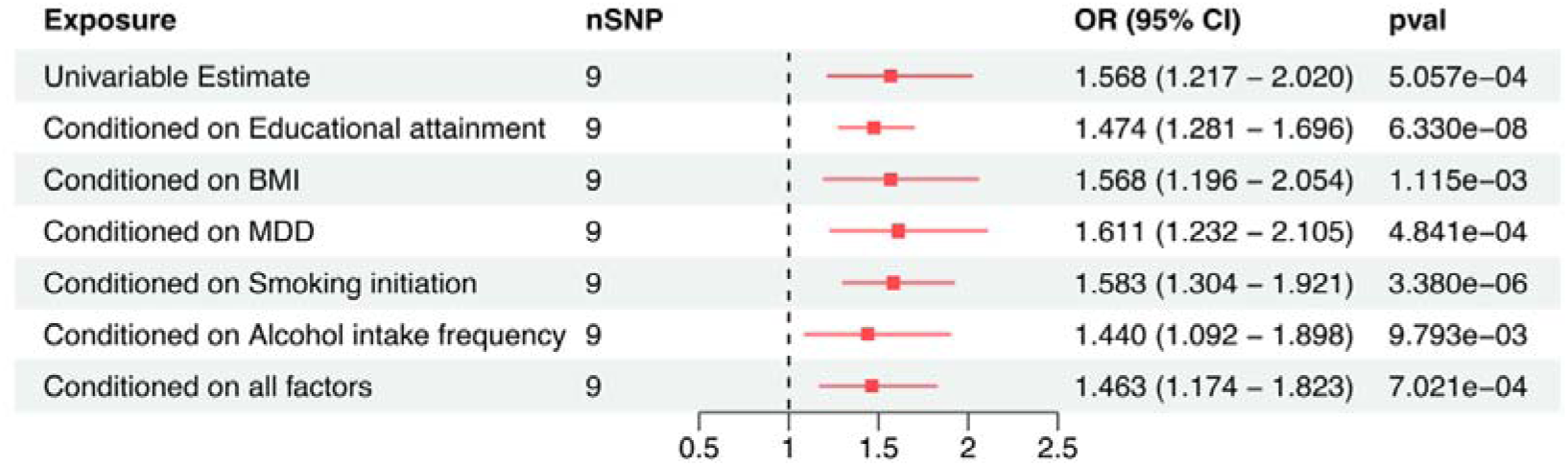
Genetically predicted association of CM and OA after adjusting for confounders Abbreviations: CI: Confidence Interval; OR: Odds Ratio; pvallJ<lJ0.05 was considered statistically significant.

Therefore, we considered BMI, MDD, smoking, alcohol, and education as potential mediators in the relationship between CM and OA. These variables were included in the mediation MR analysis to explore their mediating roles further.

#### Mediation MR

The mediation MR analysis provided substantial evidence for the mediating role of various factors in the association between CM and OA (Supplementary material Table S3&S4). The analysis indicated significant positive associations between CM and several potential mediators, including BMI (Beta= 0.148, 95% CI= 0.053 to 0.243, P=5.72e-05, MoE=0.6), MDD (Beta= 0.679, 95% CI= 0.427 to 0.932, P=1.35e-07, MoE=0.64), smoking initiation (Beta= 0.234, 95% CI= 0.119 to 0.350, P=6.81e-05, MoE=0.62), alcohol intake frequency (Beta= 0.230, 95% CI= 0.116 to 0.343, P=7.56e-05, MoE=0.63), and educational attainment (Beta= −0.706, 95% CI= −1.124 to −0.288, P=0.000545, MoE=0.54).

Furthermore, significant associations were also found between these mediators and OA. BMI was strongly associated with OA (Beta= 0.404, 95% CI= 0.376 to 0.432, P=1.46e-175, MoE=0.78). Positive associations were observed for MDD (Beta= 0.127, 95% CI= 0.064 to 0.190, P=4.81e-05, MoE=0.66), smoking initiation (Beta= 0.172, 95% CI= 0.120 to 0.224, P=3.78e-12, MoE=0.56), alcohol intake frequency (Beta= 0.168, 95% CI= 0.114 to 0.223, P=1.21e-09, MoE=0.79), and educational attainment (Beta= −0.047, 95% CI= −0.057 to −0.037, P=5.83e-20, MoE=0.52).

The mediation effect of BMI was 0.070 (95% CI: 0.030 to 0.089), accounting for 13.46% of the total effect, indicating a significant role in linking CM to OA. MDD’s mediation effect was 0.043 (95% CI: 0.033 to 0.140), responsible for 8.27% of the total effect, highlighting its critical impact in this pathway. Smoking initiation’s mediation effect was 0.061 (95% CI: 0.017 to 0.063), contributing 11.73% to the total effect. Alcohol intake frequency’s mediation effect was 0.156 (95% CI: 0.016 to 0.062), accounting for 30.00% of the total effect. Educational attainment’s mediation effect was 0.132 (95% CI: 0.013 to 0.054), playing a role in 26.23% of the total effect.

These findings underscore the multifactorial nature of the relationship between CM and OA, with BMI and MDD being particularly influential mediators (Figure 5). The substantial direct and indirect effects observed in this analysis highlight the complex pathways through which early-life adversities influence later health outcomes.

**Figure 5:**
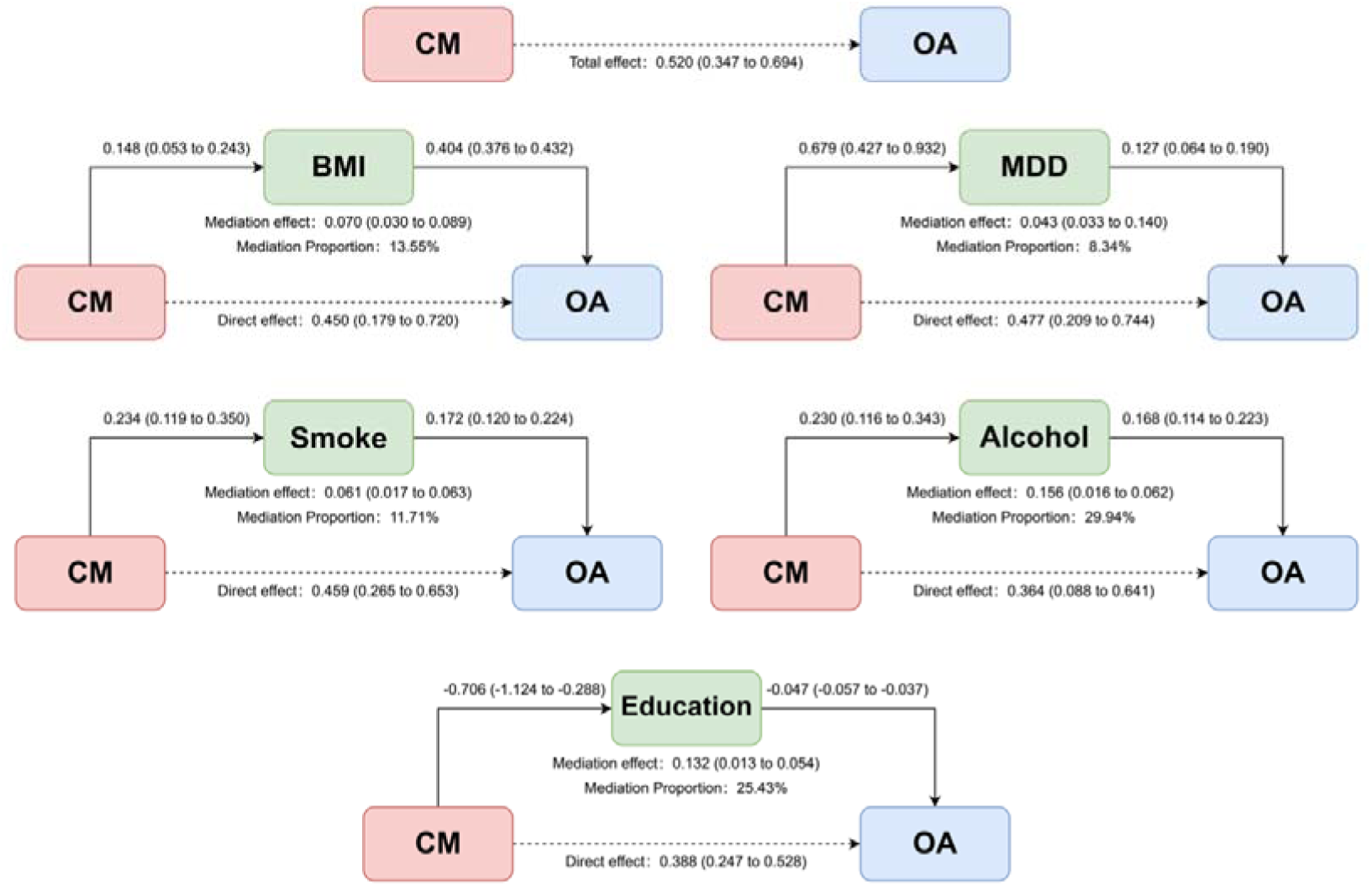
Representation of the mediation analysis showing BMI, MDD, education, and alcohol as mediators between CM and OA. Abbreviations: BMI: Body mass index; MDD: Major Depressive Disorder; Smoke: Smoking initiation; Alcohol: Alcohol intake frequency; Education: Educational attainment (years of education)

## Discussion

Our study suggested a causal effect of CM on OA that a history of CM increases the risk of OA. In this study, we found that each unit increase in CM measure raised the odds of developing OA by 68%. Multivariable MR analysis confirmed that the CM-OA relationship is significant even after adjusting for BMI, MDD, smoking, alcohol intake, and education. Mediation MR analysis identified that 13.55% of the CM-OA association is due to BMI, 8.34% to MDD, 11.71% to smoking, 29.94% to alcohol intake, and 25.43% to educational attainment. These findings highlight the multifaceted nature of the CM-OA relationship, mediated by physical health, mental health, lifestyle, and socioeconomic factors.

Our results support previous research in this area linking CM with OA. In 2009, Fuller-Thomson et al. reported that childhood physical abuse significantly increased the likelihood of developing OA in adulthood, even when controlling for various risk factors such as age, sex, race, and socioeconomic status^(19)^. Li et al. (2017) highlighted that individuals with a history of CM exhibited impaired insulin sensitivity and higher inflammatory markers, contributing to an increased risk of pre-diabetes and subsequent health complications, including OA^(40)^. Klumparendt et al. (2019) demonstrated that CM is associated with significant emotional and psychological impairments, such as increased stress and depression, which can exacerbate the risk of developing OA through inflammatory pathways^(41)^. Young-Wolff et al. (2010) have shown that individuals with a history of CM and concurrent alcohol use are at a higher risk for developing chronic health conditions, including OA, due to compounded effects of stress, inflammation, and unhealthy lifestyle choices^(42)^. Gill et al. (2021) demonstrated higher education levels are associated with lower rates of smoking and obesity, both of which are significant risk factors for OA^(43)^.

Our findings suggested intervention strategies for individuals with a history of CM should target multiple mediators because of the multifactorial nature of the CM-OA relationship. Weight management programs are essential, as increased body weight is a significant risk factor for OA. Studies show that childhood obesity causally contributes to OA, emphasizing the need for weight management. Specific strategies include dietary modifications, increased physical activity, pharmacotherapy, and, in cases of severe obesity, bariatric surgery, which has been shown to significantly reduce OA symptoms and improve metabolic health^(44, 45)^. Mental health support can help alleviate the psychological effects of CM, such as MDD and stress, which contribute to OA development. Access to mental health services, including cognitive behavioral therapy (CBT), mindfulness-based stress reduction (MBSR), and support groups, significantly improve mental well-being and reduce the psychological burden associated with CM, thereby lowering the risk of developing OA^(6, 46)^. Smoking cessation initiatives are crucial since smoking exacerbates inflammation and joint damage, further increasing the likelihood of OA in individuals with a history of CM. Programs offering support and resources for quitting smoking, including nicotine replacement therapy (NRT), behavioral interventions, and pharmacotherapy, can effectively reduce this risk^(47)^. Similarly, alcohol reduction programs are vital as excessive alcohol consumption has been linked to increased OA risk; interventions focusing on reducing alcohol intake, through education and support groups, can help mitigate this risk, especially in individuals with CM^(48)^. Educational interventions can indirectly reduce OA risk by promoting healthier lifestyles and reducing socioeconomic disparities. Education improves health literacy, enabling individuals to make informed decisions about their health, such as engaging in regular physical activity and maintaining a balanced diet^(49)^. Addressing these mediators not only helps in reducing the likelihood of developing OA but also promotes overall health and well-being. This holistic approach is essential for effectively managing and preventing OA in this vulnerable population. Our findings indicate that CM is an independent risk factor for OA, necessitating targeted risk management strategies. Weight management programs targeting BMI, mental health support for alleviating MDD and stress, smoking cessation initiatives, alcohol reduction programs, and educational interventions are all crucial components of a comprehensive intervention strategy. These measures address the multifactorial nature of the CM-OA relationship, highlighting the need for a holistic approach to mitigate the risk of OA and enhance overall health outcomes in individuals with a history of CM

Our study utilizes a robust quasi-experimental approach based on high-quality GWAS data from international consortia and employs a range of sensitivity analyses to support the main findings. However, we acknowledge several limitations. First, our findings rely on specific assumptions of the instrumental variable approach, including the absence of bias due to horizontal pleiotropy, where genetic variants influence multiple traits through different pathways. Although our results remained robust after sensitivity analyses, horizontal pleiotropy cannot be entirely ruled out due to incomplete understanding of the biological action of many included SNPs. Second, confounding effects from population stratification and dynastic effects may introduce bias. Third, the unavailability of sex-specific GWASs prevented us from conducting sex-disaggregated analyses, despite known sex dimorphism in CM and mediators such as BMI, MDD, alcohol consumption, education, and smoking. Similarly, age-specific effects were not specifically investigated. Fourth, the GWASs of our phenotypes predominantly included individuals of European ancestry, limiting generalizability. Fifth, while MR minimizes certain biases inherent in observational studies, heterogeneity in phenotype measurement can still affect our results. Differences in how phenotypes like CM and OA are measured across datasets can introduce variability and misclassification. For example, self-reported CM might vary in accuracy compared to clinical diagnoses or structured interviews, leading to measurement error. Similarly, OA diagnosed through hospital records may not capture all cases, particularly milder or undiagnosed cases, potentially biasing the result. These discrepancies can dilute true associations or create spurious ones. Sixth, the number of IVs available for our MR analysis was limited. This constraint can reduce the power of the MR analysis and potentially introduce weak instrument bias, affecting the reliability of our causal inferences. Finally, the use of large biobanks and consortia data may introduce selection bias, as participants may not be representative of the general population, affecting the generalizability of the findings.

Future research must incorporate more ethnically diverse cohorts to improve the applicability of the results. Specifically, sex-disaggregated and age-specific analyses are necessary to understand how these demographic factors modulate the CM-OA relationship. Investigating other potential mediators such as diet, physical activity, and genetic predispositions will provide a more comprehensive approach to intervention. Intervention trials targeting identified mediators should be conducted to assess their effectiveness in reducing OA risk, and research findings should be translated into practical guidelines and policies for clinical and community settings. Collaboration with healthcare providers, policymakers, and community organizations will be crucial in developing and disseminating effective intervention strategies. Addressing these future directions will enhance our understanding of the complex relationship between CM and OA and help develop more effective strategies to reduce the burden of OA in this vulnerable population.

In summary, by using a quasi-experimental approach that strengthens causal inference, this study provides robust evidence of the contribution of CM to OA and the mediating roles of BMI, MDD, smoking, alcohol intake frequency, and educational attainment. Targeted management of these mediators in individuals with a history of CM can help reduce the risk of OA and mitigate the global burden of the disease.

## Declarations

### Ethics approval and consent to participate

Not applicable

### Consent for publication

Not applicable

### Availability of data and materials

The datasets generated and analyzed during the current study are publicly available in various repositories. Specifically, the Childhood maltreatment GWAS data was derived from Apollo and can be accessed at https://www.repository.cam.ac.uk/items/70267011-0d79-4c57-87dd-3069ced8bed0. The Discovery phase osteoarthritis GWAS dataset was obtained from the Genetics of Osteoarthritis (GO) Consortium GWAS and is available at https://msk.hugeamp.org/downloads.html. Additionally, the Replication phase osteoarthritis GWAS dataset was accessed through the MRC-IEU consortium (Accessed ID: ukb-b-14486). Other GWAS datasets utilized in this study, all accessed through the MRC-IEU consortium, include Body mass index (BMI) GWAS data (Accessed ID: ukb-b-19953), Smoking initiation GWAS data (Accessed ID: ieu-b-4877), Major depressive disorder (MDD) GWAS data (Accessed ID: ieu-a-1188), Alcohol intake frequency GWAS data (Accessed ID: ukb-b-5779), and Educational attainment GWAS data (Accessed ID: ebi-a-GCST90029013). All Mediator datasets and Replication phase osteoarthritis GWAS data can be accessed at https://gwas.mrcieu.ac.uk/.

### Authors’ contributions

Zekai Yu: Supervision, Methodology, Writing - Review & Editing; Jianyu Zhu: Conceptualization, Methodology, Software, Writing - Original Draft;

## Acknowledgments

We gratefully acknowledge the GWAS consortia for providing the high-quality summary data that made this study possible.

